# Economic impact of the first wave of the COVID-19 pandemic on acute care hospitals in Japan

**DOI:** 10.1101/2020.11.09.20228791

**Authors:** Jung-ho Shin, Daisuke Takada, Tetsuji Morishita, Hueiru Lin, Seiko Bun, Emi Teraoka, Takuya Okuno, Hisashi Itoshima, Hiroyuki Nagano, Kenji Kishimoto, Hiromi Segawa, Yuka Asami, Takuya Higuchi, Kenta Minato, Susumu Kunisawa, Yuichi Imanaka

**Affiliations:** Department of Healthcare Economics and Quality Management, Graduate School of Medicine, Kyoto University, Kyoto, Japan; Department of Pharmacy, National Center for Child Health and Development Hospital, Tokyo, Japan

## Abstract

**Background:** In response to the coronavirus diseases 2019 (COVID-19) pandemic, the Japanese government declared a state of emergency on April 7, 2020. Six days earlier, the Japan Surgical Society had recommended postponing elective surgical procedures. Along with the growing public fear of COVID-19, hospital visits in Japan decreased.

**Methods:** Using claims data from the Quality Indicator/Improvement Project (QIP) database, this study aimed to clarify the impact of the first wave of the pandemic, considered to be from March to May 2020, on case volume and claimed hospital charges in acute care hospitals during this period. To make year-over-year comparisons, we considered cases from July 2018 to June 2020.

**Results:** A total of 2,739,878 inpatient and 53,479,658 outpatient cases from 195 hospitals were included. In the year-over-year comparisons, total claimed hospital charges decreased in April, May, June 2020 by 7%, 14%, and 5%, respectively, compared to the same months in 2019. Our results also showed that per-case hospital charges increased during this period, possibly to compensate for the reduced case volumes. Regression results indicated that the hospital charges in April and May 2020 decreased by 6.3% for hospitals without COVID-19 patients. For hospitals with COVID-19 patients, there was an additional decrease in proportion with the length of hospital stay of COVID-19 patients including suspected cases. The mean additional decrease per COVID-19 patients was estimated to 5.5 million JPY.

**Conclusion:** It is suggested that the hospitals treating COVID-19 patients were negatively incentivized.

## Introduction

The coronavirus disease 2019 (COVID-19) has caused serious financial difficulties for hospitals [1-3]. One of the reasons for the decrease in hospital visits is that hospitals are postponing or canceling scheduled visits, especially for surgical procedures [1,4]. Public fear of COVID-19 is also considered to be one of the reasons for the decrease [5,6]. In Japan, the first wave of COVID-19 struck in March 2020 [7]. In response, the Japanese government declared a state of emergency on April 7 [8]. The Japan Surgical Society had already announced recommendations for surgery for COVID-19 patients or those suspected of having COVID-19 on April 1, and urged the postponement of elective surgical procedures [9]. One survey has reported 5.8%, 11.4%, 3.1% decreases in the incomes of Japanese hospitals in April, May, and June 2020, respectively, compared to their incomes in the same months of the previous year [10].

On April 18, 2020, the Ministry of Health, Labour and Welfare (MHLW) doubled payments of emergency rooms and intensive care units for severe COVID-19 patients [11]. In addition, a compensation program was implemented on June 16 for securing beds for COVID-19 patients, providing 41,000 to 301,000 JPY per bed per day for moderate to severe COVID-19 patients and from 16,000 to 52,000 JPY per bed per day for mild patients [12]. However, the actual impact of the COVID-19 pandemic on the number of hospital visits and the amount of claimed hospital charges during the first wave of the pandemic has not been comprehensively examined. To fill this gap, this study aimed to clarify the impact of the first wave of the pandemic on case volumes and claimed hospital charges among Japanese acute care hospitals during the COVID-19 state of emergency.

## Methods

### Data source

We used Diagnosis Procedure Combination (DPC) data from the Quality Indicator/Improvement Project (QIP) database. The QIP database contains DPC data submitted by acute care hospitals that voluntarily participate in the project.

The DPC/per-diem payment system (PDPS) is a Japanese prospective payment system applied to acute care hospitals. In 2018, 1,730 hospitals were using the DPC/PDPS, accounting for 54% (482,618/891,872) of all general beds in Japanese hospitals [13,14]. The DPC data consist of a number of data files, including Forms 1, 3, and 4, and files D, E, F, H, and K [15]. For this study, we used forms 1 and 3, and files D, E, and F. Form 1 contains discharge summaries that include characteristics of patients, such as, sex, age, height, and weight, as well as the patients’ main diagnosis, the reason for admission, the most and second-most medical-resource-intensive diagnoses, up to 10 comorbidities, and 10 complications using the codes from the International Classification of Diseases, Tenth Revision (ICD-10). Form 3 contains hospital information, such as numbers of beds, and qualifications for special claims, such as emergency rooms and intensive care units. The D files contain information on claimed charges for both DPC/per-diem payments and fee-for-services amounts for services not covered by the per-diem payment. We extracted claimed hospital charges for each hospitalization from the D files. Files E and F contain information on all medical services, medications, and equipment for both inpatients and outpatients. These files also include the number of earned points according to the tariff of the National Health Insurance System, with one point corresponding to 10 JPY. Thus, we were able to calculate claimed charges for outpatients from the E and F files.

### Study population

We included all cases including both inpatients and outpatients, for hospitals that provided DPC data from Forms 1 and 3, files D, E, and F continuously over the study period, which extended from July 1, 2018, to June 30, 2020. Inpatient cases with a date of discharge and outpatient cases with a visit date within the study period were included. We excluded data for hospitals for which more than 10% of data monthly D file data could not be linked to Form 1 due to mismatches in patient IDs. Hospital charges for patients at hospitals with no more than 10% mismatched IDs for the D files in a month were imputed as the mean charges in the same month and the same hospital.

### Descriptive statistics

We presented descriptive statistics for the hospitals and patients. For hospitals, we presented the number of general beds; that is, hospital beds that are not psychiatric, infectious diseases, or tuberculosis beds according to the Japanese classification of hospital beds. We also computed the mean, standard deviation (SD), median, and the first and third quartiles (1Q and 3Q) for the monthly numbers of inpatient and outpatient cases and the claimed hospital charges. For inpatient and outpatient cases, we presented the number of males and the mean, median, and 1Q and 3Q for the ages and the claimed hospital charges. For inpatient cases, we also presented the number of urgent admissions and admissions with surgery, and the mean, median, and 1Q and 3Q for the length of hospital stay (LOS).

### Year-over-year comparisons

We conducted year-over-year comparisons of the monthly number of cases and claimed hospital charges. The year-over-year comparisons for inpatients cases were done according to the following categories: scheduled/urgent admissions, numbers of beds, with/without COVID-19 patients, and age strata of patients. We also compared the changes in hospital charges per case and LOS.

### Statistical analyses

We fitted linear regression models in which the dependent variable was the mean of monthly hospital charges in April and May 2020. The candidates of predictors included numbers of hospital beds (three categories: ≤199, 200–499, ≥500), mean numbers of inpatients and outpatients, mean claimed hospital charges (including both inpatients and outpatients), mean numbers of urgent hospitalizations, mean numbers of hospitalizations for surgery, and total numbers and cumulative LOS of confirmed COVID-19 patients and suspected COVID-19 patients in April and May 2020. All of the mean values indicated here were calculated using 21-month data covering the period from July 2018 through March 2020. On the other hand, the numbers and LOS of COVID-19 patients were defined as those for April and May 2020, the same period as the period used for the dependent variable, so that the potential association between these variables (i.e., the number and LOS of COVID-19 patients and hospital charges) could be evaluated. We conducted a stepwise variable selection procedure using Schwarz information criteria to choose the best set of predictors. Model 1 included just a predictor—mean hospital charges from July 2018 to March 2020. The predictors in model 2 were based on the results of the stepwise selection process, as described above.

All statistical analyses were performed using SAS software version 9.4 (SAS Institute Inc., Cary, NC, USA).

### Ethical considerations

This study was conducted in accordance with the Ethical Guidelines for Medical and Health Research Involving Human Subjects of the Ministry of Health, Labour and Welfare, Japan. The Ethics Committee, Graduate School of Medicine, Kyoto University approved the study (approval number: R0135).

## Results

### Study population

S1 Fig shows the flow of selection for the study population. A total of 195 hospitals were included, with 2,739,878 inpatient and 53,479,658 outpatient cases. It was found that 33,400 inpatient cases (1.2%) in Form 1 of the DPC data could not be matched to cases in the D file; hospital charges for these cases were imputed as the mean charges in the same hospital in the same month. For the 4,680 hospital-months in the study, the mean number of cases with imputed hospital charges was 7.14 (SD: 10.3, median: 3, interquartile range: 1 to 9).

Table 1 shows the characteristics of the hospitals. The mean monthly numbers of inpatient and outpatient cases were 585 and 11,427, respectively. During the state of emergency, these numbers decreased to 480 and 9,156, respectively (Table S1 in Supplementary Appendix).

**Table 1.**
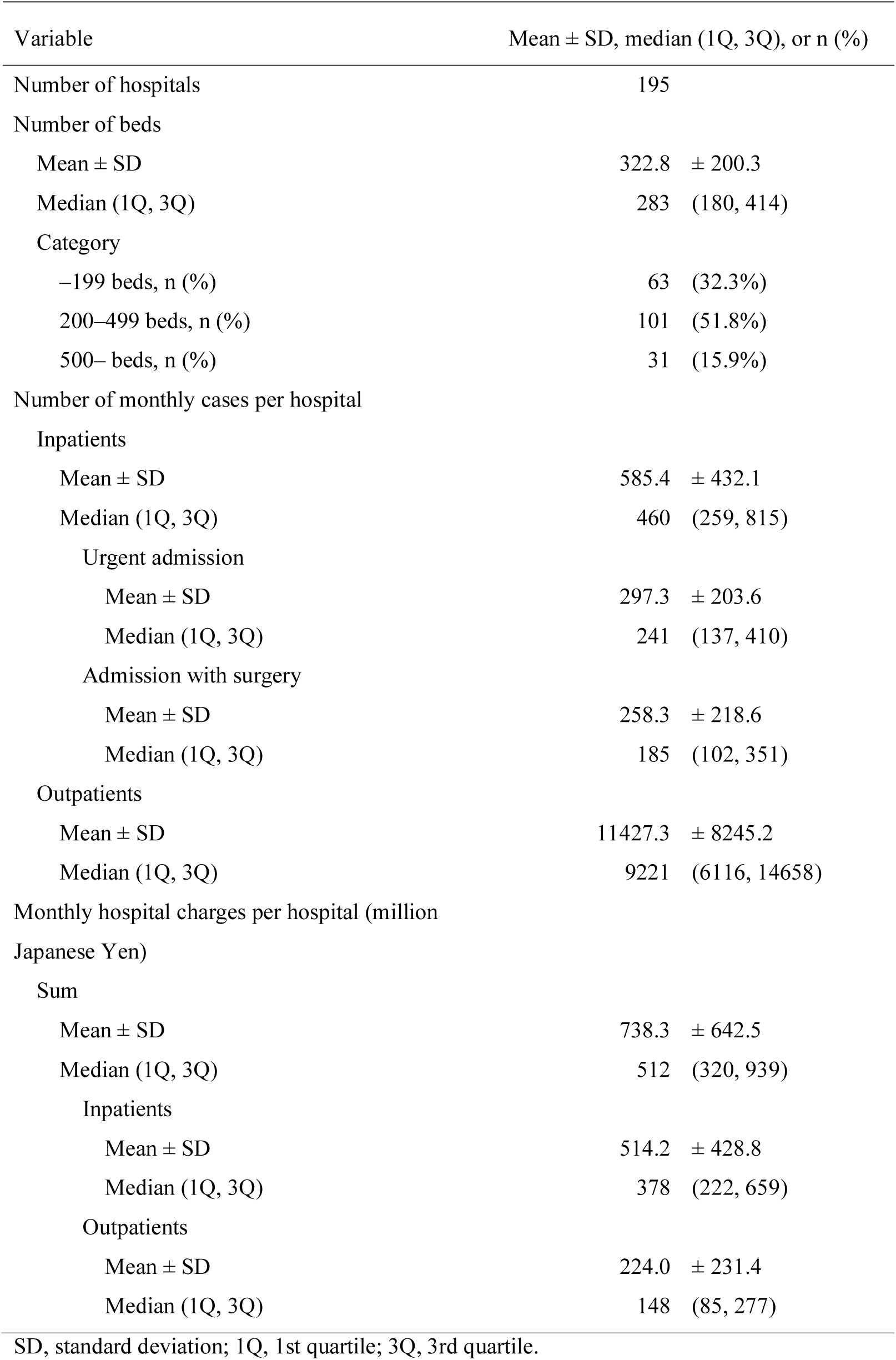
Characteristics of hospitals for the study

| Variable | Mean ± SD, median (1Q, 3Q), or n (%) |
| --- | --- |
| Number of hospitals | 195 |
| Number of beds |  |
| Mean ± SD | 322.8 ± 200.3 |
| Median (1Q, 3Q) | 283 (180, 414) |
| Category |  |
| –199 beds, n (%) | 63 (32.3%) |
| 200–499 beds, n (%) | 101 (51.8%) |
| 500– beds, n (%) | 31 (15.9%) |
| Number of monthly cases per hospital |  |
| Inpatients |  |
| Mean ± SD | 585.4 ± 432.1 |
| Median (1Q, 3Q) | 460 (259, 815) |
| Urgent admission |  |
| Mean ± SD | 297.3 ± 203.6 |
| Median (1Q, 3Q) | 241 (137, 410) |
| Admission with surgery |  |
| Mean ± SD | 258.3 ± 218.6 |
| Median (1Q, 3Q) | 185 (102, 351) |
| Outpatients |  |
| Mean ± SD | 11427.3 ± 8245.2 |
| Median (1Q, 3Q) | 9221 (6116, 14658) |
| Monthly hospital charges per hospital (million Japanese Yen) |  |
| Sum |  |
| Mean ± SD | 738.3 ± 642.5 |
| Median (1Q, 3Q) | 512 (320, 939) |
| Inpatients |  |
| Mean ± SD | 514.2 ± 428.8 |
| Median (1Q, 3Q) | 378 (222, 659) |
| Outpatients |  |
| Mean ± SD | 224.0 ± 231.4 |
| Median (1Q, 3Q) | 148 (85, 277) |

SD, standard deviation; 1Q, 1st quartile; 3Q, 3rd quartile.

S2 Table gives the characteristics of the inpatient and outpatient cases, including the mean ages—63.5 years for inpatients and 60.9 years for outpatients. Also shown are the mean hospital charges for inpatients (878,352 JPY) and outpatients (19,606 JPY). During the pandemic, these mean hospital charges increased to 973,584 JPY for inpatients and 21,747 JPY for outpatients (S3 Table in Supplementary Appendix).

### Year-over-year comparisons

S2 Fig shows the year-over-year comparison of the monthly sum of hospital charges for inpatients and outpatients. These charges decreased in April through June 2020, with May’s decrease of 100 million JPY per hospital being especially prominent. In percentage terms, hospital charges for April, May, and June 2020 decreased by 7%, 14%, and 5%, respectively, compared to the same months in 2019. S3 Fig separates the year-over-year comparisons for inpatients and outpatients. The trends appear to be quite similar. Interestingly, S3 Fig shows an approximately 10 percentage point difference between the decrease in the number of cases and the decrease in hospital charges, with the percentage decrease in hospital charges being the smaller of the two.

S4 Fig shows comparisons of the number of cases and the claimed hospitalization charges categorized by scheduled and urgent admissions. The decreases from March 2020 did not differ substantially between elective and urgent admissions. However, for admissions involving surgery, the decreases in hospital charges and the number of admissions for elective surgery was considerable, whereas the decrease in urgent admissions for surgery was less than 10% compared to the previous year (S5 Fig in Supplementary Appendix).

S6 Fig shows year-over-year comparisons of the number of cases and hospital charges according to the number of hospital beds. The patterns of these decreases did not differ among hospitals with different numbers of beds.

S7 and S8 Figs show year-over-year comparisons of the number of cases and hospital charges according to the patients’ ages. The percentage decreases were more prominent in patients aged 0–17 years.

S9 Fig shows comparisons of the number of cases and hospital charges for inpatients in hospitals with and without COVID-19 patients in 2020. As can be seen, the slopes of the decreases were steeper for hospitals with COVID-19 patients than for those without COVID-19 patients. The lowest percentages in the year-over-year comparisons of hospital charges in hospitals with and without COVID-19 patients were 86% and 91%, respectively. However, the decreases in the number of cases and charges for outpatients did not differ substantially between these two hospital categories (S10 Fig in Supplementary Appendix).

S11 and S12 Figs show hospital charges per case and mean LOS for the hospitalization cases. Fig 1 summarizes the year-over-year ratios of monthly case numbers, hospital charges, per-case hospital charges, and LOS. As indicated, the number of cases decreased from March and rebounded in June. However, per-case hospital charges for both inpatients and outpatients increased from March, although per-diem hospital charges for inpatients remained the same throughout the year.

**Fig 1.**
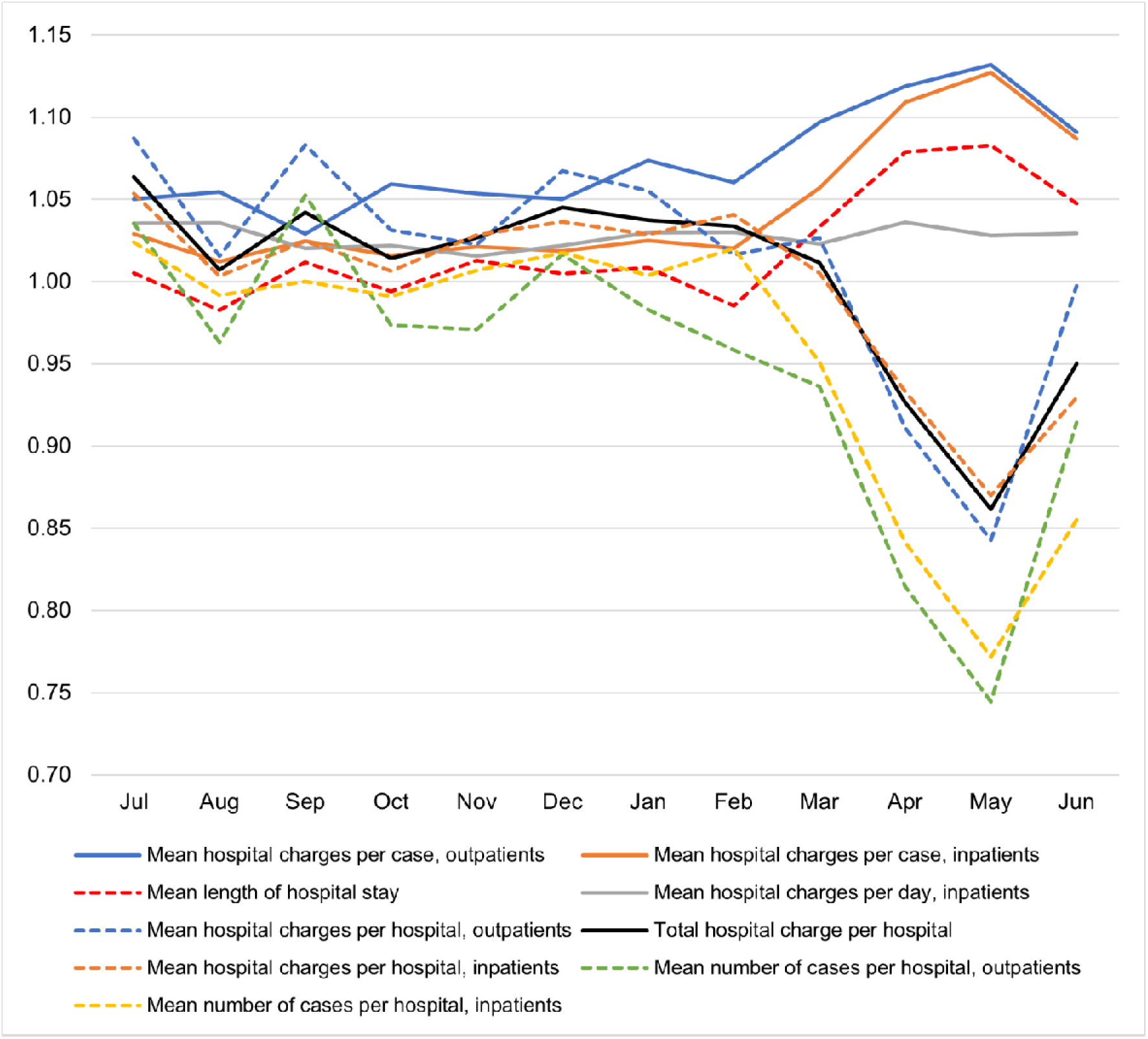
Year-over-year ratios of the number of cases, hospital charges, per-case hospital charges, and length of hospital stay.

### COVID-19 cases

S4 Table shows the number of hospitals with COVID-19 patients, including suspected COVID-19 patients, and the number and cumulative LOS of these cases. The number of confirmed cases peaked in April 2020, whereas the number of suspected cases peaked in May 2020. In June 2020, the number of suspected cases was almost 20 times more than that of confirmed cases. Table 2 shows the LOS of COVID-19 cases including suspected cases. The mean LOS of confirmed COVID-19 cases in May and June 2020 was longer than that of the earlier cases.

**Table 2.**
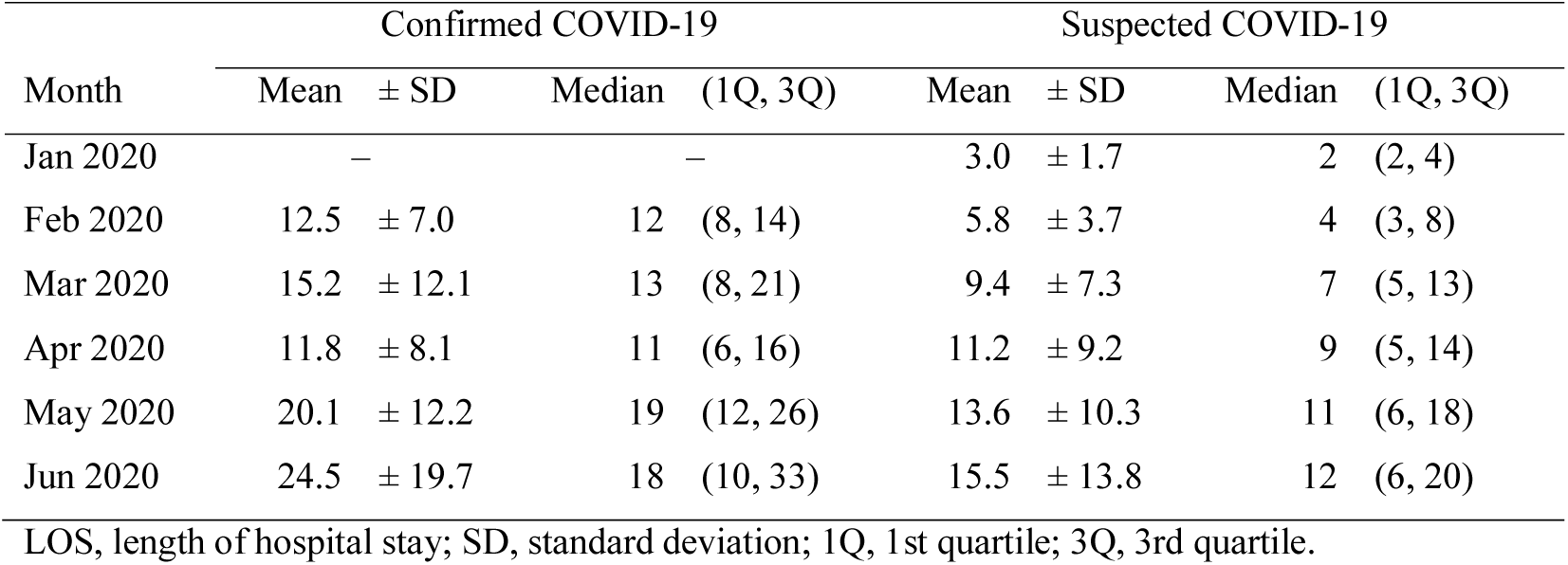
The LOS of COVID-19 cases (day)

| Month | Confirmed COVID-19 |  |  |  | Suspected COVID-19 |  |  |  |
| --- | --- | --- | --- | --- | --- | --- | --- | --- |
|  | Mean | ± SD | Median | (1Q, 3Q) | Mean | ± SD | Median | (1Q, 3Q) |
| Jan 2020 | – |  | – |  | 3.0 | ± 1.7 | 2 | (2, 4) |
| Feb 2020 | 12.5 | ± 7.0 | 12 | (8, 14) | 5.8 | ± 3.7 | 4 | (3, 8) |
| Mar 2020 | 15.2 | ± 12.1 | 13 | (8, 21) | 9.4 | ± 7.3 | 7 | (5, 13) |
| Apr 2020 | 11.8 | ± 8.1 | 11 | (6, 16) | 11.2 | ± 9.2 | 9 | (5, 14) |
| May 2020 | 20.1 | ± 12.2 | 19 | (12, 26) | 13.6 | ± 10.3 | 11 | (6, 18) |
| Jun 2020 | 24.5 | ± 19.7 | 18 | (10, 33) | 15.5 | ± 13.8 | 12 | (6, 20) |

LOS, length of hospital stay; SD, standard deviation; 1Q, 1st quartile; 3Q, 3rd quartile.

### Results of regression analyses

Table 3 shows the results of our regression analyses. In model 1, the regression coefficient of monthly claimed hospital charges was 0.892 (95% confidence interval, CI: 0.874 to 0.909, *p*<0.001); hospital charges decreased by 11% in April and May 2020. The adjusted R^2^ for model 1 was 0.981. The stepwise variable selections included three variables, the mean total hospital charges of the previous 21-month period, and the cumulative LOS of confirmed and suspected COVID-19 patients in April and May 2020, which we incorporated as predictors in model 2. With the inclusion of these two variables, the regression coefficient of monthly claimed hospital charges was changed to 0.937 (95% CI: 0.920 to 0.954, *p*<0.001), while the coefficient of the LOS of confirmed COVID-19 patients was −0.327 (95% CI: −0.400 to −0.255, *p*<0.001). Also, the coefficient of the LOS of suspected COVID-19 patients was −0.053 (95% CI: −0.086 to −0.019, *p*=0.002). The adjusted R^2^ for model 2 was 0.987. The result showed that the decrease in claimed hospital charges for hospitals without COVID-19 patients was 6.3% of the charges in the previous 21-month period. However, the claimed charges for hospitals with COVID-19 patients decreased by an additional 327,000 JPY per one day of LOS for confirmed COVID-19 patients and 53,000 JPY per day for suspected COVID-19 patients. The mean LOS of confirmed and suspected COVID-19 patients between April and June 2020 was 16.9 days and 14.1 days, respectively. Thus, the mean decrease per confirmed COVID-19 patient was 5.5 million JPY, whereas the decrease per suspected COVID-19 patient was 747,300 JPY.

**Table 3.** Results of the regression analyses

| Variable | Model 1 |  |  | Model 2 |  |  |
| --- | --- | --- | --- | --- | --- | --- |
|  | Estimate | (95% CI) | P | Estimate | (95% CI) | P |
| Monthly claimed charges before the pandemic | 0.892 | (0.874 to 0.909) | <.001 | 0.937 | (0.920 to 0.954) | <.001 |
| LOS of confirmed COVID-19 patients* | – | – | – | –0.327 | (–0.400 to –0.255) | <.001 |
| LOS of suspected COVID-19 patients* | – | – | – | –0.053 | (–0.086 to –0.019) | 0.002 |
| Adjusted $R^2$ | 0.981 | | | 0.987 | | |

CI, confidence interval; LOS, length of hospital stay.
\*Sum of LOS for each hospital in April and May 2020.

## Discussion

We analyzed claims data from July 2018 to June 2020 for both inpatients and outpatients at 195 hospitals in Japan. Our year-over-year comparisons showed decreases in both the number of cases and claimed hospital charges from March 2020 compared to the previous year. The results of our regression analyses showed that the impact of the COVID-19 pandemic on the decrease in hospital charges was more prominent for hospitals that had COVID-19 patients.

In Japan, the daily number of confirmed COVID-19 patients exceeded 100 for the first time on March 27, 2020, and the number of those requiring hospitalization increased exponentially from the end of March [16]. The Japanese Government declared a state of emergency on Apr 7, 2020 for seven of Japan’s prefectures, then expanded the declaration to cover all 47 prefectures on April 16, 2020 [8,17]. In response to the guidance of the American College of Surgeons [18], the Japan Surgical Society had already announced its recommendations for surgery involving patients with COVID-19 or those suspected of having COVID-19 on April 1, 2020 [9]. The nationwide state of emergency was lifted on May 25, 2020 [19]. Our results showed that decreases in the number of cases and claimed charges, which began in March 2020, accelerated in April and May, then reversed beginning in June 2020.

The MHLW announced a temporary increase in payments for severe COVID-19 patients on April 18, 2020 [11]. Payments for emergency rooms and intensive care units were doubled, and the limitation on the number of days for which the fees could be claimed was extended from 14 days to 21 or 35 days, depending on the status of the patient [11]. These payments were tripled on May 26 [20]. In our data, there were only 11 such claims for these doubled payments in six hospitals (data not shown). This indicates that the increase in payments for severe COVID-19 patients did not compensate for the decrease in income of hospitals with COVID-19 patients. Our regression analyses showed that the LOS of COVID-19 patients, including suspected patients, was associated with an additional decrease in claimed hospital charges. Other hospital-level variables were not selected in the stepwise process. One of the reasons for this might be that the mean of claimed charges during the previous 21-month period was such a strong predictor that other hospital-level variables were not particularly effective for predicting claimed charges in April and May 2020.

Our results suggested that the decrease in claimed charges was in proportion with the LOS of COVID-19 patients regardless of their severity. This implies hospitals treated COVID-19 patients were negatively incentivized. The current raise for payments was focused on severe COVID-19 patients, although the proportion of severe cases was not very high in Japan. Moreover, our results showed that the elective cases decreased more in April and May 2020 and returned slower in June than urgent cases. This suggested that the current compensation program that focused on severe COVID-19 patients was not sufficiently effective for the Japanese situation. Thus, more effective financial rescue policies are urgently needed for acute care hospitals. Coronavirus Aid, Relief, and Economic Security (CARES) Act Provider Relief Fund [21], which includes general distribution in proportion with gross revenue of each healthcare provider, could be one of such examples. The financial rescue policy should include compensations for overall decreases in hospital incomes as well as for COVID-19 patients.

Our results showed that the number of visits, especially elective ones, were not recovered sufficiently in June 2020, after the lift of the state of emergency. Although the reason is unclear, “a climate of fear” was suggested as one of the reasons [5]. It could change the patterns of hospital visits permanently. Under universal health coverage and “free access” with a weak gatekeeping function in Japan, people with mild diseases used to visit acute care hospitals. Policymakers should prepare for the change of paradigm in the health care system. Focus more on prevention and management, not treatment, would make a more resilient health care system.

There are several limitations to the study. First, we did not adjust for the severity of patient cases. Thus, the increase in per-case hospital charges might be due to the changes in the severity of the patient’s condition during the pandemic. Second, our data were from voluntarily participating hospitals. For external generalizability, further studies are warranted. Third, we analyzed claimed hospital charges only. Other payments, including the compensation program for securing beds for COVID-19 patients [12], were not included in the analyses.

Despite these limitations, our study analyzed more than 2 million inpatient and 53 million outpatient cases, including 187,190 inpatient and 3,570,926 outpatient cases for April and May 2020, when the national state of emergency was in force. After the declaration of the state of emergency, the numbers and claimed charges dramatically decreased in both inpatient and outpatients of acute care hospitals in Japan. Moreover, there were additional decreases in claimed charges in proportion to the number of COVID-19 patients. The raised payments, which are only for the severe COVID-19 patients, did not compensate for the decrease in revenue. Eventually, hospitals and staff were negatively incentivized, which may lead to future damage to the health care system. In addition to the reform of the reimbursement system, a financial rescue policy is urgently needed for acute care hospitals.

## Data Availability

The datasets generated during and/or analyzed during the current study are available from the corresponding author on reasonable request.

## Supporting information

S1 Fig. Flow showing the selection of hospitals for the study

S1 Table. Characteristics of hospitals for the study (comparisons of before and after the state of emergency)

S2 Table. Characteristics of the study population by inpatients and outpatients a. Inpatients

S3 Table. Characteristics of the study population by inpatients and outpatients (comparisons of before and after the state of emergency)

S2 Fig. Year-over-year comparison of the monthly sum of total hospital charges

S3 Fig. Year-over-year comparisons of the number of cases and claimed hospital charges

S4 Fig. Comparisons of the number of cases and the claimed hospitalization charges categorized by elective and urgent admissions

S5 Fig. Comparisons of the number of cases and the claimed hospitalization charges categorized by elective and urgent admissions involving surgery

S6 Fig. Comparisons of the number of cases and hospital charges according to the number of hospital beds

S7 Fig. Comparisons of the number of cases and hospital charges according to the patients’ ages

S8 Fig. Year-over-year comparisons of the number of cases and hospital charges according to the patients’ ages

S9 Fig. Year-over-year comparisons of the number of cases and hospital charges for inpatients in hospitals with and without COVID-19 patients

S10 Fig. year-over-year comparison of the monthly sum of hospital charges for inpatients and outpatients

S11 Fig. Year-over-year comparisons of hospital charges per case for the hospitalization cases

S12 Fig. Year-over-year comparisons of mean length of hospital stay for the hospitalization cases

S4 Table. The Number and cumulative LOS of COVID-19 cases

## Author contributions

Conceptualization: all authors

Methodology: J.S., D.T., and T.M.

Software: J.S., D.T., and T.M.

Validation: J.S., D.T., T.M., H.L., S.B., E.T., T.O., H.I., H.N., K.K., S.K., and Y.I.

Formal analysis: J.S., H.S., Y.A., T.H., and K.M.

Investigation: J.S., D.T., T.M., H.L., S.B., E.T., T.O., H.I., H.N., K.K., and Y.I.

Resources: S.K. and Y.I.

Data Curation: J.S. and S.K.

Writing – original draft preparation: J.S.

Writing – review and editing: all authors

Visualization: J.S.

Supervision: Y.I.

Project administration: Y.I.

Funding acquisition: Y.I.

