## Supporting information for "Economic impact of the first wave of the COVID-19 pandemic on acute care hospitals in Japan"

Contents

S1 Fig. Flow showing the selection of hospitals for the study

S1 Table. Characteristics of hospitals for the study (comparisons of before and after the state of emergency)

S2 Table. Characteristics of the study population by inpatients and outpatients

a. Inpatients

S3 Table. Characteristics of the study population by inpatients and outpatients (comparisons of before and after the state of emergency)

S2 Fig. Year-over-year comparison of the monthly sum of total hospital charges

S3 Fig. Year-over-year comparisons of the number of cases and claimed hospital charges

S4 Fig. Comparisons of the number of cases and the claimed hospitalization charges categorized by elective and urgent admissions

S5 Fig. Comparisons of the number of cases and the claimed hospitalization charges categorized by elective and urgent admissions involving surgery

S6 Fig. Comparisons of the number of cases and hospital charges according to the number of hospital beds

S7 Fig. Comparisons of the number of cases and hospital charges according to the patients’ ages

S8 Fig. Year-over-year comparisons of the number of cases and hospital charges according to the patients’ ages

S9 Fig. Year-over-year comparisons of the number of cases and hospital charges for inpatients in hospitals with and without COVID-19 patients

S10 Fig. year-over-year comparison of the monthly sum of hospital charges for inpatients and outpatients

S11 Fig. Year-over-year comparisons of hospital charges per case for the hospitalization cases

S12 Fig. Year-over-year comparisons of mean length of hospital stay for the hospitalization cases

S4 Table. The Number and cumulative LOS of COVID-19 cases

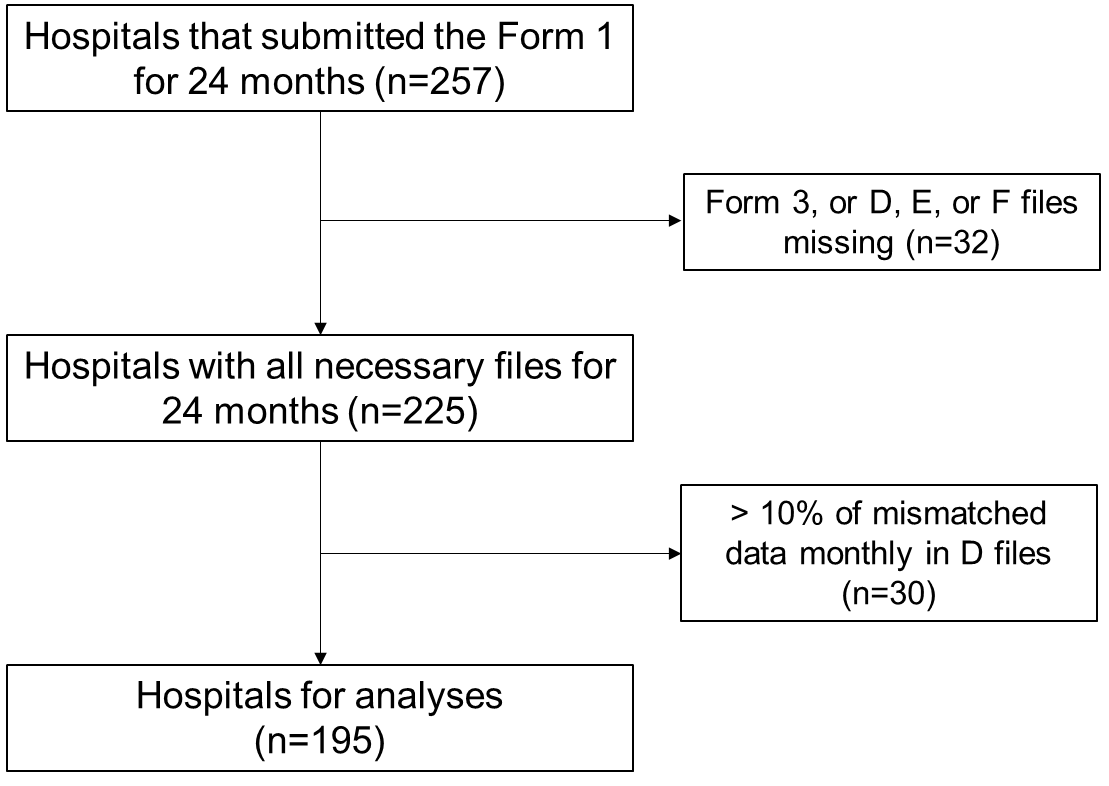

Figure S1. Flow showing the selection of hospitals for the study.

Table S1. Characteristics of hospitals for the study (comparisons of before and after the state of emergency)

| Variable | Before | | After | |
| --- | --- | --- | --- | --- |
|  | (July 2018 to March 2020) | | (April to May 2020) | |
| Number of monthly cases per hospital | | | | |
| Inpatients |  |  |  |  |
| Mean ± SD | 598.9 | ± 442.7 | 480.0 | ± 357.5 |
| Median (1Q, 3Q) | 468 | (261, 827) | 377 | (211, 656) |
| Urgent admission |  |  |  |  |
| Mean ± SD | 303.3 | ± 207.9 | 250.6 | ± 174.5 |
| Median (1Q, 3Q) | 244 | (140, 421) | 199 | (113, 342) |
| Admission with surgery |  |  |  |  |
| Mean ± SD | 263.6 | ± 223.2 | 215.4 | ± 185.5 |
| Median (1Q, 3Q) | 189 | (104, 359) | 159 | (83, 291) |
| Outpatients |  |  |  |  |
| Mean ± SD | 11685.5 | ± 8431.1 | 9156.2 | ± 6603.6 |
| Median (1Q, 3Q) | 9470 | (6294, 14971) | 7322 | (4730, 11886) |
| Monthly hospital charges per hospital (million Japanese Yen) | | | | |
| Sum of inpatients and outpatients | | |  |  |
| Mean ± SD | 746.9 | ± 650.1 | 666.4 | ± 585.3 |
| Median (1Q, 3Q) | 516 | (320, 949) | 466 | (288, 849) |
| Inpatients |  |  |  |  |
| Mean ± SD | 520.4 | ± 434.9 | 467.3 | ± 388.3 |
| Median (1Q, 3Q) | 375 | (224, 671) | 337 | (199, 593) |
| Outpatients |  |  |  |  |
| Mean ± SD | 226.5 | ± 233.0 | 199.1 | ± 214.8 |
| Median (1Q, 3Q) | 151 | (87, 282) | 132 | (71, 242) |

SD, standard deviation; 1Q, 1st quartile; 3Q, 3rd quartile.

Table S2. Characteristics of the study population by inpatients and outpatients

a. Inpatients

| Variable | Mean ± SD, median (1Q, 3Q), or n (%) | |
| --- | --- | --- |
| Number of cases | 2,739,878 |  |
| Sex |  |  |
| Male | 1,444,673 | (52.7%) |
| Age |  |  |
| Mean ± SD | 63.5 | ± 24.0 |
| Median (1Q, 3Q) | 71 | (53, 80) |
| Category |  |  |
| –17 | 215,882 | (7.9%) |
| 18–64 | 793,509 | (29.0%) |
| 65– | 1,730,487 | (63.2%) |
| Urgent admission | 1,391,213 | (50.8%) |
| Admission with surgery | 1,209,073 | (44.1%) |
| Hospital charges per case (million Japanese Yen) | |  |
| Mean ± SD | 878,352 | ± 1,182,174 |
| Median (1Q, 3Q) | 527,604 | (245142, 1055375) |
| Length of hospital stay (day) |  |  |
| Mean ± SD | 15 | ± 25.8 |
| Median (1Q, 3Q) | 8 | (4, 16) |

b. Outpatients

| Variable | Mean ± SD, median (1Q, 3Q), or n (%) | |
| --- | --- | --- |
| Number of cases | 53,479,658 |  |
| Sex |  |  |
| Male | 26,488,867 | (49.5%) |
| Age |  |  |
| Mean ± SD | 60.9 | ± 22.0 |
| Median (1Q, 3Q) | 68 | (50, 77) |
| Category |  |  |
| –17 | 3,857,731 | (7.2%) |
| 18–64 | 19,539,647 | (36.5%) |
| 65– | 30,082,280 | (56.2%) |
| Hospital charges per case (million Japanese Yen) | |  |
| Mean ± SD | 19,606 | ± 63,260 |
| Median (1Q, 3Q) | 8,060 | (3020, 20980) |

SD, standard deviation; 1Q, 1st quartile; 3Q, 3rd quartile.

Table S3. Characteristics of the study population by inpatients and outpatients (comparisons of before and after the state of emergency)

a. Inpatients

| Variable | Before | | After | |
| --- | --- | --- | --- | --- |
|  | (July 2018 to March 2020) | | (April to May 2020) | |
| N | 2,452,529 |  | 187,190 |  |
| Sex |  |  |  |  |
| Male | 1,292,266 | (52.7%) | 98,972 | (52.9%) |
| Age |  |  |  |  |
| Mean ± SD | 63.3 | ± 24.2 | 65.3 | ± 22.4 |
| Median (1Q, 3Q) | 70 | (53, 80) | 71 | (56, 81) |
| Category |  |  |  |  |
| –17 | 199,806 | (8.1%) | 10,457 | (5.6%) |
| 18–64 | 709,623 | (28.9%) | 54,781 | (29.3%) |
| 65– | 1,543,100 | (62.9%) | 121,952 | (65.1%) |
| Urgent admission | 1,241,848 | (50.6%) | 97,744 | (52.2%) |
| Admission with surgery | 1,079,599 | (44.0%) | 84,003 | (44.9%) |
| Hospital charges per case (million Japanese Yen) | | |  |  |
| Mean ± SD | 868,960 | ± 1,167,506 | 973,584 | ± 1,282,098 |
| Median (1Q, 3Q) | 520,827 | (241560, 1042586) | 597,804 | (285803, 1187393) |
| Length of hospital stay (day) | | |  |  |
| Mean ± SD | 15 | ± 25.6 | 16 | ± 28.4 |
| Median (1Q, 3Q) | 8 | (4, 16) | 9 | (4, 18) |

*(continued)*

Table S3. *(continued)*

b. Outpatients

| Variable | Before | | After | |
| --- | --- | --- | --- | --- |
|  | (July 2018 to March 2020) | | (April to May 2020) | |
| N | 47,851,971 |  | 3,570,926 |  |
| Sex |  |  |  |  |
| Male | 23,647,550 | (49.4%) | 1,819,307 | (50.9%) |
| Age |  |  |  |  |
| Mean ± SD | 60.9 | ± 0.0 | 60.9 | ± 0.0 |
| Median (1Q, 3Q) | 68 | (0, 0) | 68 | (0, 0) |
| Category |  |  |  |  |
| –17 | 3,565,043 | (7.5%) | 179,368 | (5.0%) |
| 18–64 | 17,490,040 | (36.6%) | 1,303,849 | (36.5%) |
| 65– | 26,796,888 | (56.0%) | 2,087,709 | (58.5%) |
| Hospital charges per case (million Japanese Yen) | | | |  |
| Mean ± SD | 19,384 | ± 61,777 | 21,747 | ± 75,932 |
| Median (1Q, 3Q) | 8,020 | (3010, 20800) | 8,450 | (2980, 23220) |

SD, standard deviation; 1Q, 1st quartile; 3Q, 3rd quartile.

Mean of total hospital charges per hospital (million JPY)

|  | 2018–19 | 2019–20 |
| --- | --- | --- |
| Jul | 737.711 | 784.753 |
| Aug | 764.093 | 769.493 |
| Sep | 696.071 | 725.276 |
| Oct | 763.967 | 774.855 |
| Nov | 733.558 | 752.984 |
| Dec | 773.802 | 808.696 |
| Jan | 683.54 | 709.062 |
| Feb | 708.08 | 731.818 |
| Mar | 766.113 | 774.885 |
| Apr | 756.247 | 700.613 |
| May | 733.908 | 632.225 |
| Jun | 736.73 | 699.881 |

Mean hospital charges per hospital, year-over-year ratio

|  | 全医療費 |
| --- | --- |
| Jul | 1.06377 |
| Aug | 1.00707 |
| Sep | 1.04196 |
| Oct | 1.01425 |
| Nov | 1.02648 |
| Dec | 1.0451 |
| Jan | 1.03734 |
| Feb | 1.03352 |
| Mar | 1.01145 |
| Apr | 0.926434 |
| May | 0.86145 |
| Jun | 0.949982 |

Figure S2. Year-over-year comparison of the monthly sum of total hospital charges.

|  | Hospital charges, outpatients | Hospital charges, inpatients | Number of cases, outpatients | Number of cases, inpatients |
| --- | --- | --- | --- | --- |
| Jul | 1.08732 | 1.0536 | 1.03523 | 1.02403 |
| Aug | 1.01554 | 1.00344 | 0.963165 | 0.991508 |
| Sep | 1.08309 | 1.02456 | 1.05263 | 0.999902 |
| Oct | 1.03132 | 1.00658 | 0.973603 | 0.990934 |
| Nov | 1.02256 | 1.02819 | 0.970735 | 1.00689 |
| Dec | 1.06744 | 1.03631 | 1.01638 | 1.01751 |
| Jan | 1.05521 | 1.02871 | 0.982854 | 1.00382 |
| Feb | 1.01621 | 1.04091 | 0.958438 | 1.02028 |
| Mar | 1.02672 | 1.00511 | 0.936134 | 0.950878 |
| Apr | 0.910955 | 0.933115 | 0.814317 | 0.841318 |
| May | 0.84267 | 0.869811 | 0.744592 | 0.771759 |
| Jun | 0.997306 | 0.92949 | 0.914272 | 0.855124 |

Figure S3. Year-over-year comparisons of the number of cases and claimed hospital charges.

Figure S4. Comparisons of the number of cases and the claimed hospitalization charges categorized by elective and urgent admissions.

Hospital charges, ratio

|  | all | elective | urgent |
| --- | --- | --- | --- |
| Jul | 1.0536 | 1.05435 | 1.05296 |
| Aug | 1.00344 | 1.01813 | 0.990976 |
| Sep | 1.02456 | 1.02859 | 1.02119 |
| Oct | 1.00658 | 1.00929 | 1.00435 |
| Nov | 1.02819 | 1.01486 | 1.03968 |
| Dec | 1.03631 | 1.0456 | 1.02844 |
| Jan | 1.02871 | 1.05 | 1.0144 |
| Feb | 1.04091 | 1.05539 | 1.02918 |
| Mar | 1.00511 | 0.996225 | 1.01267 |
| Apr | 0.933115 | 0.933856 | 0.932502 |
| May | 0.869811 | 0.853702 | 0.881897 |
| Jun | 0.92949 | 0.87935 | 0.973157 |

Number of cases, ratio

|  | all | elective | urgent |
| --- | --- | --- | --- |
| Jul | 1.05966 | 1.04934 | 1.00003 |
| Aug | 1.00867 | 1.00312 | 0.980316 |
| Sep | 1.00645 | 0.992016 | 1.00744 |
| Oct | 1.00061 | 0.996893 | 0.985145 |
| Nov | 1.00936 | 1.00291 | 1.01099 |
| Dec | 1.02806 | 1.0218 | 1.01336 |
| Jan | 1.03733 | 1.04015 | 0.974069 |
| Feb | 1.03847 | 1.03299 | 1.00738 |
| Mar | 0.97623 | 0.948977 | 0.952881 |
| Apr | 0.888981 | 0.855447 | 0.827526 |
| May | 0.796873 | 0.74104 | 0.798843 |
| Jun | 0.868463 | 0.814369 | 0.897351 |

Hospital charges (million JPY)

|  | 2018–19 (all) | 2019–20 (all) | 2018–19 (elective) | 2019–20 (elective) | 2018–19 (urgent) | 2019–20 (urgent) |
| --- | --- | --- | --- | --- | --- | --- |
| Jul | 507.488 | 534.691 | 234.034 | 246.755 | 273.454 | 287.936 |
| Aug | 526.753 | 528.563 | 241.743 | 246.125 | 285.01 | 282.438 |
| Sep | 481.756 | 493.588 | 219.383 | 225.654 | 262.374 | 267.934 |
| Oct | 519.108 | 522.523 | 234.757 | 236.937 | 284.35 | 285.586 |
| Nov | 503.172 | 517.356 | 232.929 | 236.39 | 270.242 | 280.966 |
| Dec | 547.063 | 566.929 | 250.946 | 262.389 | 296.117 | 304.54 |
| Jan | 453.905 | 466.934 | 182.355 | 191.473 | 271.55 | 275.462 |
| Feb | 488.833 | 508.831 | 218.826 | 230.946 | 270.007 | 277.884 |
| Mar | 533.249 | 535.975 | 245.061 | 244.136 | 288.188 | 291.84 |
| Apr | 520.225 | 485.43 | 235.672 | 220.084 | 284.553 | 265.346 |
| May | 500.109 | 435 | 214.373 | 183.011 | 285.736 | 251.989 |
| Jun | 506.321 | 470.62 | 235.692 | 207.256 | 270.629 | 263.364 |

Number of cases

|  | 2018–19 (all) | 2019–20 (all) | 2018–19 (elective) | 2019–20 (elective) | 2018–19 (urgent) | 2019–20 (urgent) |
| --- | --- | --- | --- | --- | --- | --- |
| Jul | 602.177 | 616.646 | 293.061 | 307.52 | 309.116 | 309.126 |
| Aug | 629.247 | 623.904 | 308.773 | 309.737 | 320.475 | 314.167 |
| Sep | 566.894 | 566.838 | 277.066 | 274.854 | 289.828 | 291.985 |
| Oct | 600.52 | 595.076 | 295.889 | 294.97 | 304.631 | 300.106 |
| Nov | 588.662 | 592.717 | 298.727 | 299.596 | 289.934 | 293.121 |
| Dec | 623.823 | 634.747 | 306.939 | 313.631 | 316.884 | 321.116 |
| Jan | 538.874 | 540.934 | 242.636 | 252.379 | 296.237 | 288.556 |
| Feb | 554.227 | 565.465 | 279.051 | 288.258 | 275.177 | 277.207 |
| Mar | 606.505 | 576.712 | 311.207 | 295.328 | 295.298 | 281.384 |
| Apr | 599.288 | 504.192 | 296.035 | 253.242 | 303.253 | 250.949 |
| May | 571.697 | 441.212 | 267.874 | 198.505 | 303.823 | 242.707 |
| Jun | 591.556 | 505.854 | 301.02 | 245.141 | 290.535 | 260.712 |

Figure S5. Comparisons of the number of cases and the claimed hospitalization charges categorized by elective and urgent admissions involving surgery.

Hospital charges, ratio

|  | all | elective | urgent |
| --- | --- | --- | --- |
| Jul | 1.06927 | 1.06387 | 1.07651 |
| Aug | 1.0096 | 1.02225 | 0.992508 |
| Sep | 1.02396 | 1.0285 | 1.01793 |
| Oct | 1.02136 | 1.01356 | 1.03146 |
| Nov | 1.01168 | 1.00859 | 1.01582 |
| Dec | 1.03525 | 1.04722 | 1.01953 |
| Jan | 1.0459 | 1.05297 | 1.03839 |
| Feb | 1.06062 | 1.06233 | 1.05845 |
| Mar | 1.01332 | 0.997295 | 1.03438 |
| Apr | 0.94985 | 0.934207 | 0.9699 |
| May | 0.878347 | 0.839652 | 0.923083 |
| Jun | 0.931504 | 0.855397 | 1.03415 |

Number of cases, ratio

|  | all | elective | urgent |
| --- | --- | --- | --- |
| Jul | 1.05966 | 1.05967 | 1.05962 |
| Aug | 1.00867 | 1.01681 | 0.989135 |
| Sep | 1.00645 | 1.00635 | 1.00667 |
| Oct | 1.00061 | 1.00093 | 0.999873 |
| Nov | 1.00936 | 1.01417 | 0.997795 |
| Dec | 1.02806 | 1.03396 | 1.01452 |
| Jan | 1.03733 | 1.04643 | 1.01855 |
| Feb | 1.03847 | 1.04164 | 1.03075 |
| Mar | 0.97623 | 0.964979 | 1.00424 |
| Apr | 0.888981 | 0.870933 | 0.931021 |
| May | 0.796873 | 0.740346 | 0.917127 |
| Jun | 0.868463 | 0.803521 | 1.02875 |

Hospital charges (million JPY)

|  | 2018–19 (all) | 2019–20 (all) | 2018–19 (elective) | 2019–20 (elective) | 2018–19 (urgent) | 2019–20 (urgent) |
| --- | --- | --- | --- | --- | --- | --- |
| Jul | 298.748 | 319.442 | 171.208 | 182.144 | 127.539 | 137.298 |
| Aug | 308.967 | 311.932 | 177.541 | 181.492 | 131.425 | 130.441 |
| Sep | 280.543 | 287.264 | 159.972 | 164.532 | 120.571 | 122.732 |
| Oct | 302.206 | 308.66 | 170.621 | 172.935 | 131.585 | 135.725 |
| Nov | 300.911 | 304.427 | 172.216 | 173.695 | 128.695 | 130.731 |
| Dec | 328.677 | 340.264 | 186.619 | 195.431 | 142.058 | 144.833 |
| Jan | 254.673 | 266.362 | 131.146 | 138.093 | 123.527 | 128.269 |
| Feb | 285.872 | 303.201 | 159.821 | 169.784 | 126.05 | 133.418 |
| Mar | 317.489 | 321.718 | 180.292 | 179.805 | 137.197 | 141.913 |
| Apr | 305.957 | 290.613 | 171.873 | 160.565 | 134.084 | 130.048 |
| May | 286.094 | 251.29 | 153.404 | 128.806 | 132.69 | 122.484 |
| Jun | 301.407 | 280.762 | 173.077 | 148.049 | 128.331 | 132.713 |

Number of cases

|  | 2018–19 (all) | 2019–20 (all) | 2018–19 (elective) | 2019–20 (elective) | 2018–19 (urgent) | 2019–20 (urgent) |
| --- | --- | --- | --- | --- | --- | --- |
| Jul | 260.897 | 276.462 | 183.908 | 194.882 | 76.9897 | 81.5795 |
| Aug | 275.774 | 278.164 | 194.595 | 197.867 | 81.1795 | 80.2974 |
| Sep | 248.21 | 249.81 | 173.59 | 174.692 | 74.6205 | 75.1179 |
| Oct | 267.641 | 267.805 | 186.903 | 187.077 | 80.7385 | 80.7282 |
| Nov | 269.026 | 271.544 | 189.938 | 192.631 | 79.0872 | 78.9128 |
| Dec | 282.764 | 290.697 | 196.933 | 203.621 | 85.8308 | 87.0769 |
| Jan | 223.513 | 231.856 | 150.549 | 157.538 | 72.9641 | 74.3179 |
| Feb | 252.092 | 261.79 | 178.703 | 186.144 | 73.3897 | 75.6462 |
| Mar | 278.303 | 271.687 | 198.564 | 191.61 | 79.7385 | 80.0769 |
| Apr | 265.836 | 236.323 | 185.99 | 161.985 | 79.8462 | 74.3385 |
| May | 244.031 | 194.462 | 166 | 122.897 | 78.0308 | 71.5641 |
| Jun | 268.503 | 233.185 | 191.082 | 153.538 | 77.4205 | 79.6462 |

Figure S6. Comparisons of the number of cases and hospital charges according to the number of hospital beds.

Hospital charges (million JPY)

|  | –199 | 200–499 | 500– |
| --- | --- | --- | --- |
| Jul | 1.04494 | 1.04791 | 1.0633 |
| Aug | 0.995199 | 1.00015 | 1.00998 |
| Sep | 1.06443 | 1.03886 | 0.995795 |
| Oct | 0.988872 | 1.01719 | 0.998411 |
| Nov | 1.02644 | 1.02249 | 1.03601 |
| Dec | 1.03106 | 1.04893 | 1.02232 |
| Jan | 1.04184 | 1.03844 | 1.01237 |
| Feb | 1.021 | 1.03103 | 1.05957 |
| Mar | 1.00425 | 1.01341 | 0.99483 |
| Apr | 0.954167 | 0.938668 | 0.920215 |
| May | 0.882007 | 0.868958 | 0.867211 |
| Jun | 0.92994 | 0.942057 | 0.91358 |

Number of cases

|  | –199 | 200–499 | 500– |
| --- | --- | --- | --- |
| Jul | 1.00394 | 1.02025 | 1.03645 |
| Aug | 0.984089 | 0.986636 | 1.00105 |
| Sep | 0.994377 | 1.00104 | 1.00017 |
| Oct | 0.977782 | 0.986368 | 1.00198 |
| Nov | 1.01282 | 0.997632 | 1.01823 |
| Dec | 0.99701 | 1.02168 | 1.01869 |
| Jan | 0.991367 | 1.00554 | 1.00565 |
| Feb | 1.00206 | 1.01423 | 1.03507 |
| Mar | 0.93352 | 0.95979 | 0.9442 |
| Apr | 0.850773 | 0.84097 | 0.838663 |
| May | 0.783241 | 0.76875 | 0.772181 |
| Jun | 0.851478 | 0.862968 | 0.845206 |

Hospital charges (million JPY)

|  | 2018-19 (–199) | 2019-20 (–199) | 2018-19 (200–499) | 2019-20 (200–499) | 2018-19 (500–) | 2019-20 (500–) |
| --- | --- | --- | --- | --- | --- | --- |
| Jul | 177.203 | 185.167 | 495.066 | 518.783 | 1268.3 | 1348.59 |
| Aug | 187.019 | 186.121 | 512.087 | 512.163 | 1315.94 | 1329.08 |
| Sep | 168.431 | 179.284 | 463.41 | 481.416 | 1224.91 | 1219.76 |
| Oct | 185.982 | 183.913 | 501.686 | 510.308 | 1303.1 | 1301.03 |
| Nov | 181.333 | 186.128 | 490.618 | 501.652 | 1246.83 | 1291.73 |
| Dec | 192.254 | 198.226 | 524.655 | 550.325 | 1394.08 | 1425.19 |
| Jan | 162.585 | 169.387 | 442.922 | 459.948 | 1125.65 | 1139.58 |
| Feb | 177.523 | 181.252 | 476.92 | 491.72 | 1207.61 | 1279.55 |
| Mar | 187.33 | 188.126 | 519.28 | 526.245 | 1333.36 | 1326.47 |
| Apr | 183.742 | 175.321 | 502.096 | 471.301 | 1313.45 | 1208.66 |
| May | 183.59 | 161.928 | 489.303 | 425.183 | 1226.96 | 1064.04 |
| Jun | 181.372 | 168.665 | 489.559 | 461.192 | 1270.31 | 1160.53 |

Number of cases

|  | 2018-19 (–199) | 2019-20 (–199) | 2018-19 (200–499) | 2019-20 (200–499) | 2018-19 (500–) | 2019-20 (500–) |
| --- | --- | --- | --- | --- | --- | --- |
| Jul | 233.714 | 234.635 | 612.594 | 625 | 1375.32 | 1425.45 |
| Aug | 241.429 | 237.587 | 639.366 | 630.822 | 1445.32 | 1446.84 |
| Sep | 220.19 | 218.952 | 571.881 | 572.475 | 1310.1 | 1310.32 |
| Oct | 233.619 | 228.429 | 607.188 | 598.911 | 1382.55 | 1385.29 |
| Nov | 227.841 | 230.762 | 597.98 | 596.564 | 1348.55 | 1373.13 |
| Dec | 244.222 | 243.492 | 623.95 | 637.475 | 1455.23 | 1482.42 |
| Jan | 209.603 | 207.794 | 547.257 | 550.287 | 1232.87 | 1239.84 |
| Feb | 215.571 | 216.016 | 558.535 | 566.485 | 1282.06 | 1327.03 |
| Mar | 232.079 | 216.651 | 608.446 | 583.98 | 1419.81 | 1340.58 |
| Apr | 228.905 | 194.746 | 602.911 | 507.03 | 1398.19 | 1172.61 |
| May | 221.444 | 173.444 | 583.228 | 448.356 | 1301.26 | 1004.81 |
| Jun | 228.175 | 194.286 | 597.317 | 515.465 | 1368.52 | 1156.68 |

Figure S7. Comparisons of the number of cases and hospital charges according to the patients’ ages.

Hospital charges, outpatients (million JPY)

|  | 2018–19 (0–17) | 2019–20 (0–17) | 2018–19 (18–64) | 2019–20 (18–64) | 2018–19 (65–) | 2019–20 (65–) |
| --- | --- | --- | --- | --- | --- | --- |
| Jul | 10.2095 | 10.5037 | 81.155 | 86.7489 | 131.051 | 144.583 |
| Aug | 11.6749 | 11.5042 | 83.6945 | 84.1182 | 133.866 | 137.175 |
| Sep | 9.80464 | 10.2783 | 75.1098 | 80.5463 | 121.989 | 133.27 |
| Oct | 10.8053 | 10.4647 | 84.6117 | 86.7748 | 141.456 | 147.053 |
| Nov | 10.419 | 10.1224 | 79.5116 | 81.115 | 132.715 | 136.431 |
| Dec | 11.7065 | 11.534 | 78.8297 | 84.0149 | 127.786 | 137.497 |
| Jan | 12.0578 | 11.2105 | 80.3719 | 84.1663 | 130.223 | 139.568 |
| Feb | 11.1018 | 9.85397 | 76.1522 | 77.1623 | 124.472 | 128.143 |
| Mar | 11.9765 | 9.78509 | 81.0059 | 83.2819 | 131.678 | 137.597 |
| Apr | 9.88917 | 6.33008 | 80.8323 | 75.244 | 137.298 | 126.141 |
| May | 9.59118 | 5.72571 | 80.7684 | 68.8556 | 135.745 | 115.951 |
| Jun | 9.52857 | 7.36371 | 80.037 | 79.0145 | 133.054 | 135.642 |

Number of cases, outpatients

|  | 2018–19 (0–17) | 2019–20 (0–17) | 2018–19 (18–64) | 2019–20 (18–64) | 2018–19 (65–) | 2019–20 (65–) |
| --- | --- | --- | --- | --- | --- | --- |
| Jul | 947.61 | 975.092 | 4408.04 | 4507.92 | 6712.11 | 7009.94 |
| Aug | 1025.03 | 980.974 | 4556.53 | 4359.84 | 6763.11 | 6549.14 |
| Sep | 793.056 | 843.128 | 4003.39 | 4151.83 | 6068.11 | 6441.36 |
| Oct | 882.005 | 841.544 | 4569.7 | 4417.29 | 7111.58 | 6972.83 |
| Nov | 834.374 | 795.446 | 4325.25 | 4173.09 | 6691.57 | 6535.84 |
| Dec | 913.415 | 893.903 | 4303.48 | 4333.55 | 6439.98 | 6620.36 |
| Jan | 919.333 | 822.8 | 4364.21 | 4222.84 | 6383.52 | 6421.38 |
| Feb | 798.246 | 729.841 | 4076.92 | 3877.21 | 6115.1 | 5926.44 |
| Mar | 926.415 | 717.846 | 4355.74 | 4080.72 | 6576.94 | 6303.14 |
| Apr | 877.641 | 480.159 | 4217.34 | 3513.81 | 6713.15 | 5621.61 |
| May | 878.077 | 439.677 | 4209.32 | 3172.6 | 6592.65 | 5084.59 |
| Jun | 886.492 | 581.128 | 4178.33 | 3824.4 | 6471.67 | 6141.96 |

Hospital charges, inpatients (million JPY)

|  | 2018–19 (0–17) | 2019–20 (0–17) | 2018–19 (18–64) | 2019–20 (18–64) | 2018–19 (65–) | 2019–20 (65–) |
| --- | --- | --- | --- | --- | --- | --- |
| Jul | 27.885 | 27.3005 | 130.41 | 135.013 | 357 | 380.603 |
| Aug | 32.1734 | 32.2401 | 139.712 | 138.827 | 362.972 | 365.628 |
| Sep | 26.7338 | 26.3255 | 126.811 | 123.601 | 335.623 | 351.255 |
| Oct | 26.3317 | 26.287 | 132.687 | 132.274 | 368.076 | 372.002 |
| Nov | 23.0971 | 26.283 | 127.669 | 129.172 | 360.147 | 369.86 |
| Dec | 28.8355 | 26.8531 | 141.154 | 142.867 | 385.49 | 405.931 |
| Jan | 22.4696 | 22.3005 | 113.247 | 111.746 | 325.171 | 340.071 |
| Feb | 22.1746 | 23.502 | 121.532 | 128.049 | 352.647 | 365.108 |
| Mar | 25.9851 | 23.6482 | 133.482 | 131.277 | 381.986 | 389.296 |
| Apr | 27.0802 | 20.2419 | 127.715 | 118.283 | 373.433 | 354.373 |
| May | 25.2276 | 18.9013 | 122.248 | 109.464 | 360.328 | 313.327 |
| Jun | 24.5434 | 21.0554 | 126.987 | 117.141 | 362.581 | 339.664 |

Number of cases, inpatients

|  | 2018–19 (0–17) | 2019–20 (0–17) | 2018–19 (18–64) | 2019–20 (18–64) | 2018–19 (65–) | 2019–20 (65–) |
| --- | --- | --- | --- | --- | --- | --- |
| Jul | 55.2974 | 54.0564 | 179.636 | 179.821 | 376.508 | 392.256 |
| Aug | 62.5026 | 60.159 | 191.497 | 190.441 | 384.928 | 382.903 |
| Sep | 49.8974 | 49.7487 | 172.313 | 165.872 | 353.405 | 359.938 |
| Oct | 48.7949 | 47.8564 | 177.723 | 175.179 | 383.241 | 381.195 |
| Nov | 44.0205 | 44.3026 | 173.615 | 172.733 | 380.082 | 384.8 |
| Dec | 50.5077 | 50.0821 | 184.379 | 185.113 | 398.533 | 409.318 |
| Jan | 43.8564 | 42.3385 | 154.154 | 154.995 | 349.154 | 351.923 |
| Feb | 39.5385 | 40.359 | 161.862 | 165.897 | 361.354 | 367.908 |
| Mar | 50.1641 | 40.3846 | 176.938 | 168.544 | 388.733 | 376.656 |
| Apr | 51.7795 | 29.0513 | 171.487 | 147.728 | 385.241 | 335.169 |
| May | 49.3692 | 24.5744 | 164.436 | 133.2 | 366.687 | 290.226 |
| Jun | 49.6308 | 28.8154 | 172.456 | 149.256 | 378.569 | 335.564 |

Figure S8. Year-over-year comparisons of the number of cases and hospital charges according to the patients’ ages.

Number of cases, year-over-year ratio

|  | 65– (outpatient) | 18–64 (outpatient) | 65– (inpatient) | 18–64 (inpatient) | 0–17 (outpatient) | 0–17 (inpatient) |
| --- | --- | --- | --- | --- | --- | --- |
| Jul | 1.04437 | 1.02266 | 1.04183 | 1.00103 | 1.029 | 0.977557 |
| Aug | 0.968362 | 0.956834 | 0.994738 | 0.994483 | 0.957019 | 0.962504 |
| Sep | 1.06151 | 1.03708 | 1.01849 | 0.96262 | 1.06314 | 0.99702 |
| Oct | 0.980489 | 0.966647 | 0.994661 | 0.985688 | 0.954126 | 0.980767 |
| Nov | 0.976727 | 0.96482 | 1.01241 | 0.99492 | 0.953344 | 1.00641 |
| Dec | 1.02801 | 1.00699 | 1.02706 | 1.00398 | 0.978638 | 0.991573 |
| Jan | 1.00593 | 0.967607 | 1.00793 | 1.00546 | 0.894996 | 0.965388 |
| Feb | 0.969148 | 0.951014 | 1.01814 | 1.02493 | 0.914306 | 1.02075 |
| Mar | 0.95837 | 0.93686 | 0.968933 | 0.952555 | 0.774864 | 0.80505 |
| Apr | 0.837402 | 0.83318 | 0.870025 | 0.861453 | 0.547102 | 0.561058 |
| May | 0.771252 | 0.753709 | 0.79148 | 0.810042 | 0.500727 | 0.497767 |
| Jun | 0.949054 | 0.915294 | 0.886401 | 0.865473 | 0.655537 | 0.580595 |

Hospital charges, year-over-year ratio

|  | 65– (outpatient) | 18–64 (outpatient) | 65– (inpatient) | 18–64 (inpatient) | 0–17 (inpatient) | 0–17 (outpatient) |
| --- | --- | --- | --- | --- | --- | --- |
| Jul | 1.10326 | 1.06893 | 1.06611 | 1.03529 | 0.979036 | 1.02882 |
| Aug | 1.02472 | 1.00506 | 1.00732 | 0.993669 | 1.00207 | 0.985378 |
| Sep | 1.09248 | 1.07238 | 1.04658 | 0.974689 | 0.984727 | 1.04831 |
| Oct | 1.03957 | 1.02556 | 1.01067 | 0.996889 | 0.998303 | 0.968484 |
| Nov | 1.028 | 1.02017 | 1.02697 | 1.01178 | 1.13794 | 0.971533 |
| Dec | 1.07599 | 1.06578 | 1.05303 | 1.01213 | 0.93125 | 0.98526 |
| Jan | 1.07176 | 1.04721 | 1.04582 | 0.986745 | 0.992475 | 0.929732 |
| Feb | 1.02949 | 1.01326 | 1.03533 | 1.05363 | 1.05986 | 0.8876 |
| Mar | 1.04495 | 1.0281 | 1.01914 | 0.983487 | 0.910069 | 0.817024 |
| Apr | 0.918741 | 0.930866 | 0.94896 | 0.926146 | 0.747482 | 0.640102 |
| May | 0.854178 | 0.852506 | 0.869562 | 0.895429 | 0.749234 | 0.596977 |
| Jun | 1.01945 | 0.987225 | 0.936797 | 0.922467 | 0.857884 | 0.772804 |

Figure S9. Year-over-year comparisons of the number of cases and hospital charges for inpatients in hospitals with and without COVID-19 patients.

Hospital charges, ratio

|  | With COVID-19 patients | Without COVID-19 patients |
| --- | --- | --- |
| Jul | 1.05479 | 1.04847 |
| Aug | 1.00362 | 1.00265 |
| Sep | 1.01944 | 1.04663 |
| Oct | 1.00149 | 1.0284 |
| Nov | 1.03272 | 1.00912 |
| Dec | 1.03521 | 1.04107 |
| Jan | 1.02967 | 1.02463 |
| Feb | 1.04374 | 1.02908 |
| Mar | 1.00082 | 1.02364 |
| Apr | 0.927518 | 0.956956 |
| May | 0.860342 | 0.909174 |
| Jun | 0.924995 | 0.948567 |

Number of cases, ratio

|  | With COVID-19 patients | Without COVID-19 patients |
| --- | --- | --- |
| Jul | 1.02767 | 1.00953 |
| Aug | 0.992182 | 0.988801 |
| Sep | 1.00355 | 0.985523 |
| Oct | 0.99434 | 0.977355 |
| Nov | 1.01206 | 0.986517 |
| Dec | 1.02086 | 1.00422 |
| Jan | 1.00723 | 0.990299 |
| Feb | 1.02426 | 1.00437 |
| Mar | 0.946514 | 0.968566 |
| Apr | 0.832427 | 0.877252 |
| May | 0.760351 | 0.817486 |
| Jun | 0.848413 | 0.882419 |

Hospital charges (million JPY)

|  | 2018–19 (with COVID-19 patients) | 2019–20 (with COVID-19 patients) | 2018–19 (without COVID-19 patients) | 2019–20 (without COVID-19 patients) |
| --- | --- | --- | --- | --- |
| Jul | 685.663 | 723.231 | 248.537 | 260.584 |
| Aug | 711.371 | 713.944 | 258.473 | 259.159 |
| Sep | 650.752 | 663.406 | 236.161 | 247.173 |
| Oct | 700.352 | 701.394 | 255.81 | 263.075 |
| Nov | 676.396 | 698.53 | 251.802 | 254.098 |
| Dec | 738.519 | 764.521 | 268.878 | 279.92 |
| Jan | 610.589 | 628.706 | 226.488 | 232.066 |
| Feb | 656.13 | 684.832 | 246.177 | 253.335 |
| Mar | 720.2 | 720.788 | 261.571 | 267.754 |
| Apr | 700.99 | 650.181 | 257.719 | 246.625 |
| May | 670.751 | 577.075 | 252.66 | 229.712 |
| Jun | 681.811 | 630.672 | 251.528 | 238.591 |

Number of cases

|  | 2018–19 (with COVID-19 patients) | 2019–20 (with COVID-19 patients) | 2018–19 (without COVID-19 patients) | 2019–20 (without COVID-19 patients) |
| --- | --- | --- | --- | --- |
| Jul | 800.798 | 822.958 | 314.947 | 317.947 |
| Aug | 838.378 | 831.824 | 326.632 | 322.974 |
| Sep | 752.261 | 754.933 | 299.026 | 294.697 |
| Oct | 798.798 | 794.277 | 313.763 | 306.658 |
| Nov | 781.252 | 790.672 | 310.342 | 306.158 |
| Dec | 829.092 | 846.387 | 327.039 | 328.421 |
| Jan | 716.437 | 721.613 | 282.118 | 279.382 |
| Feb | 737.546 | 755.437 | 289.066 | 290.329 |
| Mar | 809.445 | 766.151 | 312.684 | 302.855 |
| Apr | 799.353 | 665.403 | 309.684 | 271.671 |
| May | 761.303 | 578.857 | 297.382 | 243.105 |
| Jun | 790.017 | 670.261 | 304.158 | 268.395 |

Figure S10. year-over-year comparison of the monthly sum of hospital charges for inpatients and outpatients.

Hospital charges, inpatients

Million JPY

|  | 2018–19 | 2019–20 |
| --- | --- | --- |
| Jul | 222.415 | 241.836 |
| Aug | 229.236 | 232.798 |
| Sep | 206.903 | 224.094 |
| Oct | 236.873 | 244.293 |
| Nov | 222.646 | 227.668 |
| Dec | 218.322 | 233.046 |
| Jan | 222.652 | 234.944 |
| Feb | 211.726 | 215.159 |
| Mar | 224.661 | 230.664 |
| Apr | 228.019 | 207.715 |
| May | 226.105 | 190.532 |
| Jun | 222.62 | 222.02 |

Number of cases, outpatients

|  | 2018–19 | 2019–20 |
| --- | --- | --- |
| Jul | 12067.8 | 12493 |
| Aug | 12344.7 | 11890 |
| Sep | 10864.6 | 11436.3 |
| Oct | 12563.3 | 12231.7 |
| Nov | 11851.2 | 11504.4 |
| Dec | 11656.9 | 11847.8 |
| Jan | 11667.1 | 11467 |
| Feb | 10990.3 | 10533.5 |
| Mar | 11859.1 | 11101.7 |
| Apr | 11808.1 | 9615.57 |
| May | 11680 | 8696.87 |
| Jun | 11536.5 | 10547.5 |

Number of cases, inpatients

|  | 2018–19 | 2019–20 |
| --- | --- | --- |
| Jul | 611.441 | 626.133 |
| Aug | 638.928 | 633.503 |
| Sep | 575.615 | 575.559 |
| Oct | 609.759 | 604.231 |
| Nov | 597.718 | 601.836 |
| Dec | 633.421 | 644.513 |
| Jan | 547.164 | 549.256 |
| Feb | 562.754 | 574.164 |
| Mar | 615.836 | 585.585 |
| Apr | 608.508 | 511.949 |
| May | 580.492 | 448 |
| Jun | 600.656 | 513.636 |

Hospital charges, inpatients

Million JPY

|  | 2018–19 | 2019–20 |
| --- | --- | --- |
| Jul | 515.296 | 542.917 |
| Aug | 534.857 | 536.695 |
| Sep | 489.168 | 501.182 |
| Oct | 527.094 | 530.562 |
| Nov | 510.913 | 525.315 |
| Dec | 555.479 | 575.651 |
| Jan | 460.888 | 474.118 |
| Feb | 496.353 | 516.659 |
| Mar | 541.452 | 544.221 |
| Apr | 528.228 | 492.898 |
| May | 507.803 | 441.693 |
| Jun | 514.111 | 477.861 |

Figure S11. Year-over-year comparisons of hospital charges per case for the hospitalization cases.

Hospital charges per case, outpatients (JPY)

|  | 2018–19 | 2019–20 |
| --- | --- | --- |
| Jul | 18430.5 | 19357.8 |
| Aug | 18569.6 | 19579.4 |
| Sep | 19043.9 | 19595 |
| Oct | 18854.4 | 19972.2 |
| Nov | 18786.8 | 19789.7 |
| Dec | 18729.1 | 19669.9 |
| Jan | 19083.8 | 20488.7 |
| Feb | 19264.9 | 20426.2 |
| Mar | 18944.2 | 20777.3 |
| Apr | 19310.3 | 21602 |
| May | 19358.2 | 21908.1 |
| Jun | 19297 | 21049.5 |

Hospital charges per case, inpatients (JPY)

|  | 2018–19 | 2019–20 |
| --- | --- | --- |
| Jul | 842757 | 867095 |
| Aug | 837117 | 847187 |
| Sep | 849818 | 870774 |
| Oct | 864431 | 878079 |
| Nov | 854773 | 872855 |
| Dec | 876952 | 893157 |
| Jan | 842322 | 863200 |
| Feb | 882008 | 899846 |
| Mar | 879216 | 929364 |
| Apr | 868072 | 962788 |
| May | 874780 | 985922 |
| Jun | 855915 | 930350 |

Figure S12. Year-over-year comparisons of mean length of hospital stay for the hospitalization cases.

Mean length of hospital stay (day)

|  | 2018–19 | 2019–20 |
| --- | --- | --- |
| Jul | 14.2973 | 14.3721 |
| Aug | 14.2598 | 14.0116 |
| Sep | 14.4661 | 14.6382 |
| Oct | 14.8424 | 14.756 |
| Nov | 14.3033 | 14.4873 |
| Dec | 14.4738 | 14.5413 |
| Jan | 14.5915 | 14.717 |
| Feb | 15.1421 | 14.9195 |
| Mar | 14.8452 | 15.3385 |
| Apr | 14.5985 | 15.7458 |
| May | 15.1575 | 16.414 |
| Jun | 14.1777 | 14.8528 |

Table S4. The Number and cumulative LOS of COVID-19 cases

|  | All | | | Confirmed COVID-19 cases | | | Suspected COVID-19 cases | | |
| --- | --- | --- | --- | --- | --- | --- | --- | --- | --- |
| Month | Number of hospitals | Number of cases | Sum of LOS (day) | Number of hospitals | Number of cases | Sum of LOS (day) | Number of hospitals | Number of cases | Sum of LOS (day) |
| Jan 2020 | 3 | 3 | 9 | 0 | 0 | 0 | 3 | 3 | 9 |
| Feb 2020 | 25 | 57 | 434 | 7 | 16 | 200 | 23 | 42 | 242 |
| Mar 2020 | 70 | 338 | 3,831 | 33 | 115 | 1,746 | 62 | 225 | 2,119 |
| Apr 2020 | 147 | 1,420 | 16,282 | 93 | 513 | 6,068 | 134 | 923 | 10,343 |
| May 2020 | 160 | 2,652 | 38,912 | 87 | 451 | 9,080 | 153 | 2,209 | 29,990 |
| Jun 2020 | 150 | 2,980 | 47,438 | 49 | 150 | 3,675 | 149 | 2,834 | 43,897 |

LOS, length of hospital stay.
